# PDE5 inhibition and Alzheimer’s disease risk: a mendelian randomisation study

**DOI:** 10.1101/2024.11.16.24317338

**Authors:** Marta Alcalde-Herraiz, Benjamin Woolf, Junqing Xie, Emma Anderson, Dipender Gill, Ioanna Tzoulaki, Laura M Winchester, James Yarmolinsky, Daniel Prieto-Alhambra, Danielle Newby

**Affiliations:** Centre for Statistics in Medicine and NIHR Biomedical Research Centre Oxford, NDORMS, University of Oxford, Oxford, UK; School of Psychological Science, University of Bristol, Bristol, United Kingdom; MRC Integrative Epidemiology Unit, University of Bristol, Bristol, United Kingdom; MRC Biostatistics Unit, University of Cambridge, Cambridge, United Kingdom; Division of Psychiatry, University College of London, London, UK; Department of Epidemiology and Biostatistics, School of Public Health, Imperial College London, London, UK; MRC-PHE Centre for Environment, School of Public Health, Imperial College London, London, UK; Department of Hygiene and Epidemiology, University of Ioannina Medical School, Ioannina, Greece; Department of Psychiatry, Warneford Hospital, Oxford, UK; Department of Medical Informatics, Erasmus University Medical Centre, Rotterdam, the Netherlands

**Author notes:** Corresponding author: Prof. Daniel Prieto-Alhambra, Botnar Research Centre, Windmill Road, OX37LD, Oxford, United Kingdom.

**Keywords:** Alzheimer’s disease, Mendelian randomisation, PDE-5, Phosphodiesterase-5, dementia, sildenafil

## Abstract

**INTRODUCTION:** While preclinical studies suggest that Phosphodiesterase 5 (PDE5) inhibition may reduce cognitive impairment, findings from observational studies on whether PDE5 inhibitors reduce Alzheimer’s disease (AD) risk have been inconsistent.

**METHODS:** A two-sample *cis-*Mendelian Randomisation (MR) analysis was conducted to estimate the causal effect of PDE5 inhibition on AD risk. The analysis was performed across four different genome-wide association studies (GWAS) of AD to enhance evidence reliability through triangulation. Additionally, a sex-stratified MR analysis using data from UK Biobank was performed to assess potential sex-specific effects.

**RESULTS:** No evidence of a causal association between PDE5 inhibition and AD risk was found in the main analyses or sex-stratified analysis.

**DISCUSSION:** MR findings suggest that PDE5 inhibitors are unlikely to decrease the risk of AD. Further research is needed to thoroughly understand the impact of PDE5 inhibitors on the risk of Alzheimer’s disease.

## 1. BACKGROUND

Alzheimer’s disease (AD) is a progressive neurodegenerative disorder and the most common cause of dementia. It is characterised by the accumulation of beta-amyloid plaques and neurofibrillary tangles in the brain, leading to a gradual deterioration of cognitive function and memory^1^. With the global increase in life expectancy, AD is rapidly emerging as a significant public health thread worldwide^2^.

Recently, the Medicines and Healthcare products Regulatory Agency (MHRA) in the UK approved Lecanemab for treating adults in the early stages of AD^3^. While the treatment has been shown effective for slowing the disease progression, the National Institute for Health and Care Excellence (NICE) has not recommended its availability on the NHS, arising concerns about its cost-effectiveness^4^. There is therefore still a scarcity of drugs that can effectively treat or prevent AD.

Several studies have identified numerous modifiable risk factors that may be associated with AD^5^, offering the potential opportunity for intervention through drug repurposing strategies^6^. This strategy aims to identify new therapeutic uses for existing drugs that have already been approved. In terms of cost-effectiveness, reduced drug-development time, and lower risk of failure, repurposing presents a highly advantageous strategy. Within this context, antihypertensive and related medications have been previously highlighted as promising candidates for AD prevention^7^. Specifically, Phosphodiesterase 5 (PDE5) inhibitors have gained growing interest due to their potential neuroprotective effects^8^.

PDE5 inhibitors (i.e., sildenafil, vardenafil, tadalafil, and avanafil) are mainly used for the treatment of erectile dysfunction (ED) and pulmonary arterial hypertension (PAH)^9,10^. PDE5 is an enzyme present in smooth muscle cells whose inhibition has been shown to induce vascular smooth relaxation and vasodilatation. Simultaneously, this results in a reduction of diastolic blood pressure^11^. Some preclinical models have also highlighted the potential of PDE5 inhibitors on improving memory function^12^, although these results were not observed in subsequent clinical studies.

Conflicting results have also been reported in observational studies^13,14^, where confounding, time-related bias or reverse causation can play a key role. Alternatively, Mendelian randomisation (MR) can offer a robust approach to investigate causal relationships overcoming some of the limitations of observational research, as well as allowing for triangulation of evidence^15,16^. Specifically, drug target MR is an analytical method that uses genetic variants within or near the gene encoding the risk factor (*cis*-variants) as instruments to proxy the exposure^15,17^. Since alleles are randomly allocated during meiosis, this approach can minimise the risk of confounding that typically affects traditional observational studies, provided that all underlying MR assumptions are met.

In this study, we performed a two-sample *cis-*MR analysis to estimate the causal effect of genetically proxied PDE5 inhibition and the risk of AD. To investigate this, we have used diastolic blood pressure as a surrogate biomarker. Furthermore, we also conducted a stratified by sex MR analysis, where we used UK Biobank (UKB) to identify AD cases.

## 2. METHODS

### 2.1. Study design

We performed a two-step *cis*-Mendelian Randomisation analysis to estimate the causal effect between genetically proxied PDE5 inhibition and AD risk. We scaled the effect of PDE5 inhibition based on a surrogate biomarker -diastolic blood pressure^15,18^. We used four different genome-wide association study (GWAS) of AD, each one differing by the sample size, heritability value, and the use of by-proxy cases. Additionally, we performed a sex-stratified analysis using UKB dataset to identify AD cases. Figure 1 summarises the study design.

**Fig. 1:**
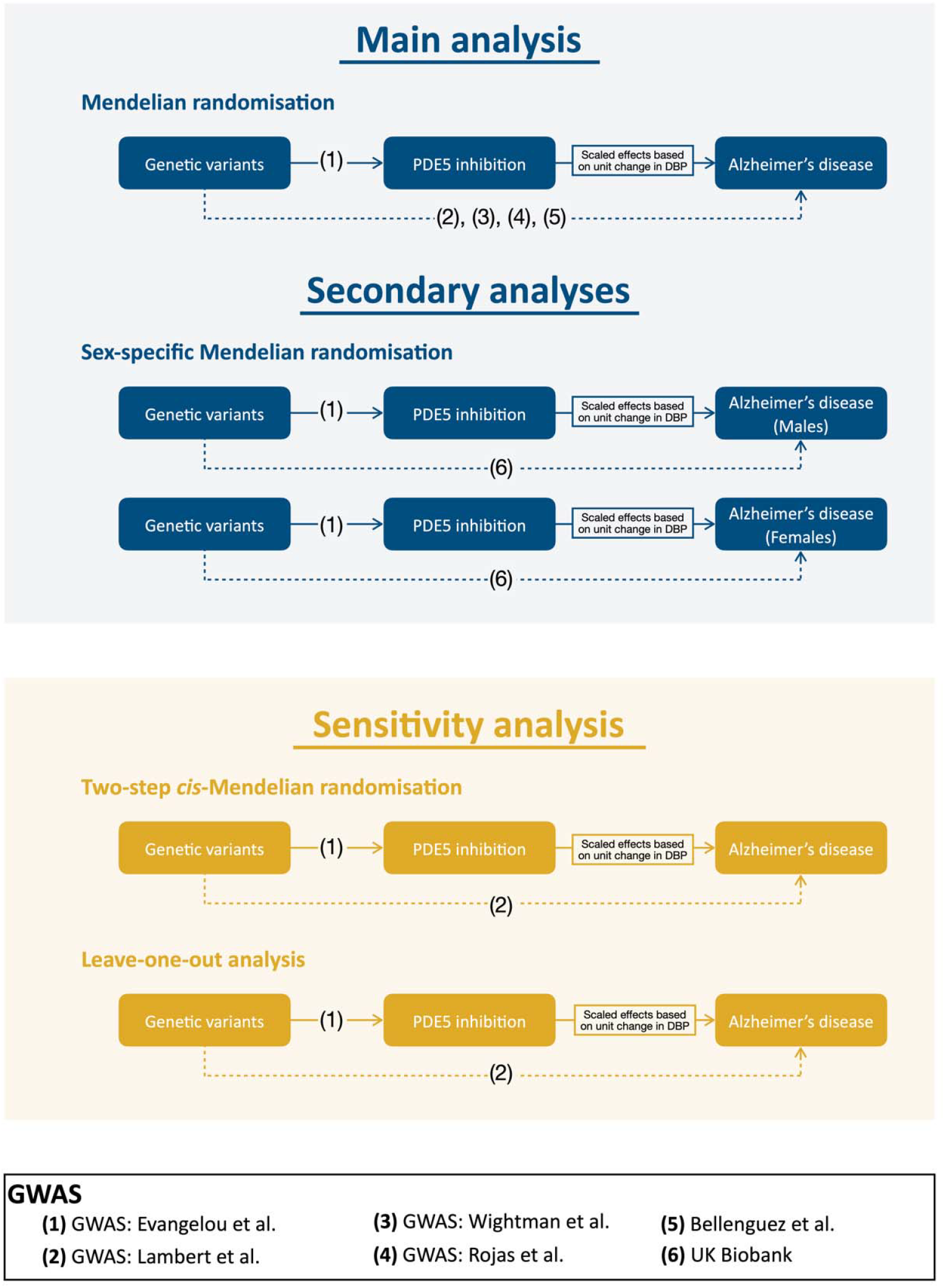
Scheme of the study design. GWAS = Genome wide association study.

### 2.2. Data sources

#### 2.2.1. Variant-risk factor estimates

Variant-exposure estimates were extracted from a GWAS of diastolic blood pressure^19^. We used diastolic blood pressure (DBP) instead of systolic blood pressure (SBP) as PDE5 inhibition is known to have a greater impact on the former^11^. Evangelou *et al*. GWAS meta-analysed 77 cohorts (n = 757,601) participating in the International Consortium for Blood Pressure Genome (ICBP)^20^ and UKB^21^. Blood pressure for each individual in the participating cohorts was measured in mmHg using either manual or automated readings. All participating cohorts adjusted for age, age^2^, sex, body mass index, and study-specific covariates. UKB cohort further corrected BP measures for those with self-reported medication use. Further information about study design, participants, and genotype quality control of this GWAS can be found in the original publication

GWAS estimates were measured in mmHg change per effect allele, and SNPs positions were reported in hg19/GRCh37 coordinates.

#### 2.2.2. Variant-outcome estimates

Genetic variant-outcome associations were derived from the AD GWAS published by Lambert *et al*.^*22*^ which conducted a two-stage meta-analysis of GWAS in individuals of European ancestry. We used estimates from stage 1, which consisted of 54,162 participants (17,008 AD cases and 37,154 controls) containing 7,055,881 genotyped and imputed variants. Further details of the databases included in this meta-analysis can be found in Supplement (Note 1 and Table 1). As secondary outcomes, we used three different AD GWAS: Rojas *et al*.^23^ (N = 409,435), Bellenguez *et al*.^*24*^ (N = 487,511) and Wightman *et al*.^*25*^ (N = 398,108). All of them included only European participants. More details about the study populations can be found in the respective original publications and in supplementary (Note 1 and Table 1).

**Table 1:**
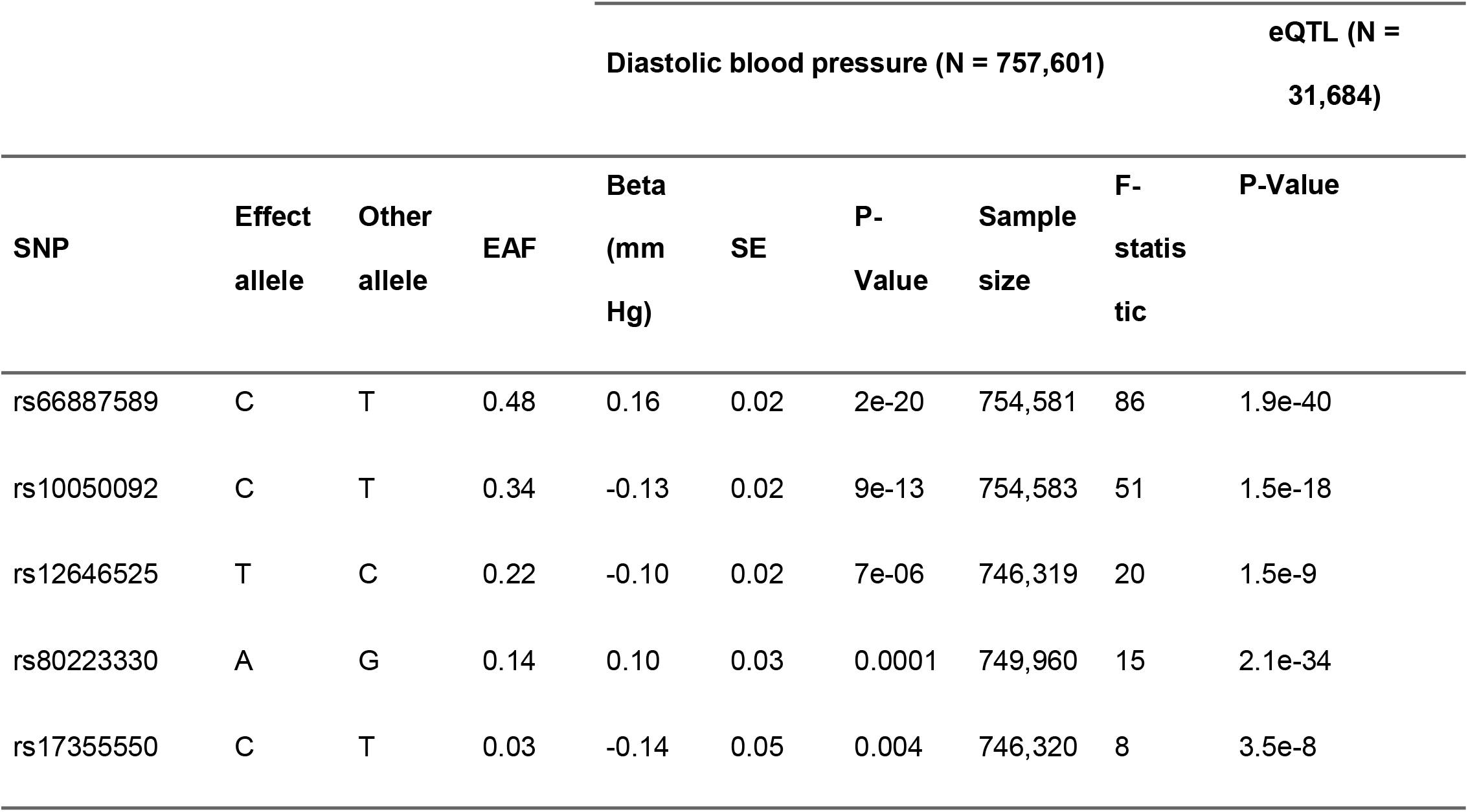
Single nucleotide polymorphisms employed as instruments for the Mendelian randomisation analysis. Note: SNP = Single nucleotide polymorphism, SE = Standard error, EAF = effect allele frequency.

We included these additional AD GWAS as secondary outcomes to validate our main study results. We did this due to the following reasons: (1) although they have bigger sample sizes, these AD GWAS have smaller SNP-based heritability value compared to Lambert AD GWAS; (2) the AD GWAS conducted by Rojas and Bellenguez contain a proportion of by-proxy-cases for AD; and, 3) Rojas and Bellenguez GWAS included UKB, as well as Evangelou blood pressure GWAS, and we wanted to avoid sample overlap as can lead to biased results in the presence of weak instruments^26^.

All GWAS estimates were reported on the *log*(OR) scale. SNP coordinates were in hg19/GRCh37 assembly, except Rojas *et al*. GWAS, which was in the hg38/GRCh38 assembly. We aligned the coordinates of this GWAS from hg38/GRCh38 to hg19/GRCh37 using LiftOver^27^.

In the sex-specific analysis, we used UKB^21^ to estimate the variant-outcome effects. UKB is a large population-based cohort study of over 500,000 participants aged 40 to 69 at recruitment (2006 and 2010). UKB collected lifestyle data and biological samples for genotyping^28^. Genotyping, performed by Affymetrix, includes 784,256 autosomal variants. Imputation was done using the Haplotype Reference Consortium and UK10K panels, covering 93 million SNPs. Health outcomes, including dementia and AD, were ascertained using validated algorithms combining baseline data and linked hospital and death records, with follow-up data until December 2022 (https://biobank.ndph.ox.ac.uk/showcase/refer.cgi?id=460). In total, there are around 10,000 of dementia cases recorded in UKB.

### 2.3. Statistical analysis

#### 2.3.1. Instrument selection

Instrumental variables used in MR must (1) be associated with the exposure, (2) not be associated with the outcome through confounding pathways, and (3) not affect the outcome except via the exposure^29^. Instrumental variables to proxy PDE5 inhibition were obtained from a previous study that also targeted PDE5 inhibition^30^. This study selected five *cis*-genetic variants within the PDE5 gene (chromosome 4, position in GRCh37/hg19 120,415,550-120,550-146). These variants were associated with PDE5 gene expression in blood at genome-wide significance (P-Value<5·10^−8^) and clumped using the P-Values of their associations with diastolic blood pressure (r^2^<0.35, window = 10,000 kilobases). Instruments from this study were validated using two positive control outcomes (erectile dysfunction and pulmonary hypertension).

These five variants extracted from Woolf *et al*. study were used as instruments for this study (Table 1). The variant with strongest association (i.e., lowest P-Value) was rs66887589 (Beta = -0.16 respect allele T, 95%CI = -0.2 to -0.12). Variant rs17355550 was the one with weakest association with DBP (Beta = -0.14 respect allele C, 95%CI = -0.23 to -0.04).

#### 2.3.2. Main analysis

Mendelian randomisation estimates were calculated using the inverse-variance weighted method accounting for correlation between variants^31^. We used the linkage disequilibrium matrix corresponding to European ancestry participants with the 1000 Genomes reference panel phase 3. Exposure alleles and outcome alleles were harmonised with respect to the linkage disequilibrium matrix. All MR estimates were reported as OR. We scaled our estimates to represent a daily 100mg dose of sildenafil, knowing that this amount decreases 5.5mmHg DBP.

#### 2.3.3. Sex-stratified analysis

As AD is more prevalent in women compared to men^32^, we repeated the main analysis separately in females and males. We used the same variant-risk factor GWAS as in the main analysis (described in section Methods/Data sources/Variant-risk factor estimates), as previous research showed that SNP effects were identical for both, the mixed and the specific sex analysis^33^. We used UKB to calculate genetic variants-AD associations separately for both females and males.

We used the algorithmically-defined dementia types to restrict our UK Biobank cohort to participants without other forms of dementia (frontotemporal, vascular, or all cause of dementia – including only those with AD-). AD cases were classified as those with an AD report, whereas AD controls were those without one. All UKB variable fields IDs used in the study can be found in Supplementary Table 2.

We estimated the effect size of the genetic instruments on the algorithmically defined AD (separately for each sex) using a logistic regression. We used age at first assessment, the genetic batch, and the first 10 genetic principal components as covariates in the model. Beta coefficients obtained from the two different regressions were used later for computing the MR estimates.

MR effects were reported as OR per 5.5mmHg decrease in DBP.

### 2.4. Sensitivity analysis

#### 2.4.1. Two-Step *cis-*Mendelian randomization

To account for any potential horizontal pleiotropic effects, we performed two-step *cis*-MR. This approach employs a two-step mediation strategy to account for potential pleiotropic pathways by adjusting variant-outcome associations^34^. Hence, this approach is used to adjust our MR estimates for any effect mediated by other related traits.

We adjusted our analysis for body mass index (BMI), as its inclusion as a covariate in the DBP GWAS can induce collider bias^35^.

#### 2.4.2. Leave-one-out analysis

To assess whether the results from the main analysis were driven by a single instrument, we performed leave-one-out analysis^36^. In this approach, one SNP from the instrumental set is removed and the estimate causal effect is re-calculated. We performed this analysis for each one of the AD GWAS included in this study.

### 2.5. Software and implementation

We used R (version 3.2) for this study. The main packages used included *TwoSampleMR*^*37*^, *TwoStepCisMR*^*34*^, *coloc*^*38*^, liftOver^27^, dplyr^39^, and ggplot2^40^. This manuscript was written according to the STROBE-MR reporting guidelines^41^.

All the analytical code can be found in the public GitHub repository: https://github.com/oxford-pharmacoepi/PDE5_AD_MendelianRandomisation

## 3. RESULTS

### 3.1. Main analysis

We found little evidence of an association between genetically proxied PDE5 inhibition and AD risk using Lambert’s *et al*. GWAS (OR = 1.00, 95% confidence interval 0.96 to 1.04, P-Value = 0.96). These results were consistent when using the other AD GWAS data (Figure 2).

**Fig. 2:**
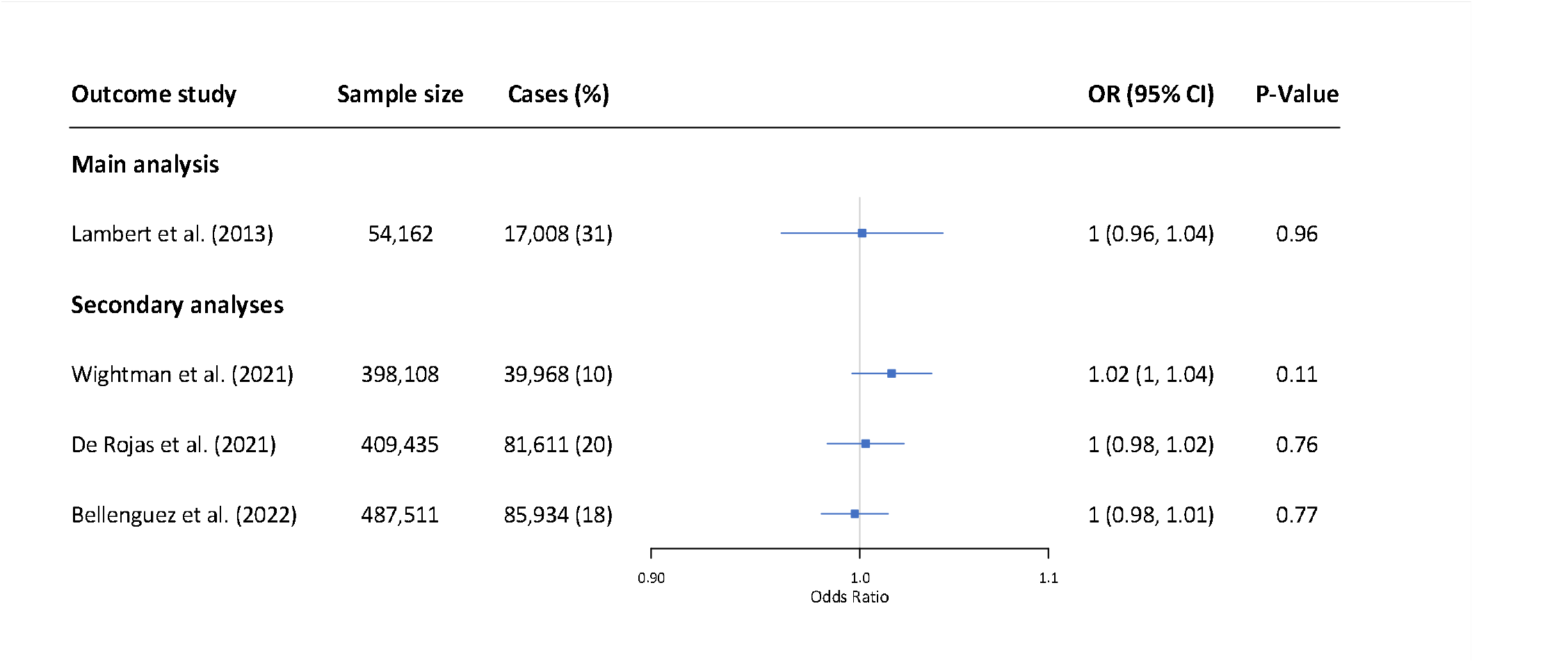
Results from the Mendelian randomization. Odds ratio is scaled to represent 100mg dose of sildenafil (which reduces 5mmHg diastolic blood pressure). Note: OR = Odds ratio, CI = Confidence interval.

Refer to Supplementary Table 3 to see the linkage disequilibrium matrix employed. Harmonised variant-risk factor and variant-outcome effects can be found in Supplementary Table 4. SNPs effect on the exposure against SNPs effects on the outcome (Lambert’s *et al*) can be found in Supplementary Figure 1.

### 3.2. Sex-stratified analysis

From the 502,230 participants in UKB, we excluded 6,153 participants because they were diagnosed with vascular dementia, frontotemporal dementia or other forms of dementia (Supplementary Figure 2). After further restricting to individuals with genotyped data, we obtained a cohort of 262,037 females with 2,030 AD cases, and a cohort of 220,352 males with 1,722 cases. Baseline characteristics of the respective cohorts can be found in Supplementary Table 5.

MR results for both sex were consistent with the main analysis (Supplementary Figure 3), showing no association between genetically proxied PDE5 inhibition and AD risk (Male: scaled OR = 1.04, 95%CI = 0.97 to 1.11, P-Value = 0.3; Female: scaled OR = 0.95, 95%CI = 0.88 to 1.03, P-Value = 0.2).

### 3.3. Sensitivity analyses

#### 3.3.1. Two-Step *cis-*Mendelian randomization

Results from the two-step *cis-*MR were consistent with the main findings (Supplementary Table 6), suggesting that BMI did not induce bias to our results.

#### 3.3.2. Leave-one-out

There was little evidence to suggest that any of the instruments was driven to the obtained results was found in the leave-one-out analysis. All results were consistent with the main findings (Supplementary Table 7).

## 4. DISCUSSION

In this study, we used drug target MR to investigate the association between genetically proxied PDE5 inhibition and the risk of AD. Using DBP to scale PDE5 inhibition, our analysis showed little evidence of an effect of PDE5 inhibition on AD risk using a variety of AD GWAS datasets. Results stratifying by sex also showed no evidence of a causal association.

PDE5 is an enzyme responsible for degrading cyclic guanosine monophosphate (cGMP)^42^, a molecule that actives protein kinase G (PKG). PKG plays a key role in the regulation of smooth muscle relaxation, particularly in blood vessels. In addition to its vascular effects, the cGMP/PKG signalling have also been shown to play a role in neuronal plasticity^43-45^. Animal models have shown the potential of sildenafil enhancing memory and cognitive function^46-49^ while having other neuroprotective effects, such as improving synaptic plasticity^50^, or reducing amyloid burden^46^. However, clinical studies results’ regarding sildenafil efficacy to enhance cognition have not been yet conclusive^12^, although it has been recently shown to improve brain blood flow^8^.

Interestingly, observational studies have also found contradictory results. In 2021, Fang *et al*.^13^ developed an endophenotype network analysis where sildenafil was identified as a candidate drug in AD. They further conducted an observational study that suggested a reduction of 69% the risk of AD among sildenafil users compared to nonusers. This result was not aligned with a new-user active comparator cohort study conducted by Desai *et al*. (2022)^14^, where they did not find any evidence of an association between sildenafil and endothelin receptor antagonist users in people with PAH. Strengths, limitations and differences between the previous two observational studies have been extensively discussed elsewhere^14,51,52^, mainly pointing to issues regarding the target population and the study design.

Our study presents genetic evidence that supports Desai *et al*.’s results. However, there are significant differences in the interpretation of both studies. First, Desai and colleagues included other dementias (vascular dementia, senile, pre-senile, or unspecified dementia) rather than AD to define the outcome, which can bias the results to the null if sildenafil has only impact on AD. In our study, we used different GWAS studies for AD, each one with different definition. For example, the main study outcome (Lambert’s *et al*.^*22*^) did include (although in small proportion) participants with mixed AD and vascular dementia (Supplementary Note 1) to define AD, as well as Bellenguez *et al*.^*24*^ definition. However, Wightman^25^ and De Rojas^23^ used a more specific definition for AD. Second, they restricted the study population to participants with PAH to minimise the risk of confounding by indication, reducing significantly the study population sample size. This can result in a lack of statistical power and generalisability to the general population, not only because of the sample size but also because, given the high mortality rate among medicated PAH patients, it can be that participants did not live long enough to develop AD. However, in our study we used Mendelian randomisation, which assumes that alleles are randomised at meiosis, we could study the impact of genetically proxied PDE5 inhibition, instrumented in a wider European population. Third, whereas they studied the impact of sildenafil usage on AD, our study did not assess that. Instead, we studied the effect of having lower levels of PDE5 in a lifelong time.

AD has been reported to be more prevalent in women^32^, whereas sildenafil is mainly prescribed to men. Hence, we performed a sex-stratified analysis using data from UKB to estimate the variant-outcome associations independently for males and females. Our findings indicated that genetically proxied PDE5 inhibition was not associated with a reduced risk of AD in either sex. However, both analyses were influenced by a significant case-control imbalance ratios, with only around 0.8% of participants being classified as cases. Hence, our sex-stratified analysis can be underpowered to detect any association.

A previous observational study conducted only in men diagnosed with ED had identified an association between the use of PDE5 inhibitors and decrease risk of AD^53^. The cohort study compared users of PDE5 inhibitors with non-users, based on the assumption that patients continued the treatment after the initial prescription. However, PDE5 inhibitors for ED are typically taken when required rather than regularly. This intermittent use challenges the assumption that participants consistently followed up with the treatment once prescribed, potentially impacting the study’s conclusions.

Our study has several strengths that enhance the robustness of our findings. Firstly, we used MR to triangulate evidence from previous observational evidence, which is known to be less susceptible to unobserved confounding and reverse causation, compared to traditional observational studies^15,17^. Second, we used instrumental variables that have been previously tested and validated using positive control outcomes, enhancing their reliability. Third, we included different AD definitions, each one prone to different bias, in the main analysis and obtained consistent evidence, ensuring the robustness of our results. Additionally, we further performed a two-step *cis-*MR to adjust for potential pleiotropic effect, where results were also consistent with the main analysis.

There are some limitations that must be considered when interpreting our results. First, by using MR, we are studying the effect of small lifelong effects of lower PDE5 levels on AD. However, PDE5 inhibitors are typically administered for a shorter time, in higher doses and at a specific point in time. Thus, the effect estimates from this study should not be interpreted as the effect of the pharmacological intervention, but as an approximation of the direction of the causal effects. Second, we only included European populations in our study (although in the sex-specific analysis we did not restrict to increase the sample size, the proportion of non-white individuals was much smaller compared to white individuals). Therefore, our results are potentially not generalisable to other populations. Third, our model assumes that the effects of PDE5 inhibition are linear across the dose-response range. Forth, although we included different AD GWAS with different sources of bias, all of them relied on clinical diagnostic rather than postmortem autopsy (which is known to be the gold standard to detect the underlying cause of dementia). Hence, heterogeneity in our outcome might influence the results, as different types of dementia can have different risk factors^54^.

Our study provides genetic evidence that PDE5 inhibitors are unlikely to decrease the risk of AD. Findings from this study have significant implications for our understanding of AD pathophysiology and the identification of PDE5 inhibitors potential therapeutic targets.

## Supporting information

STROBE-MR checklist

## 5. Data availability

All GWAS used are publicly available (see references to original publications and Supplementary Table 1). UK Biobank individual-level source data can be accessed by applying for access at http://ukbiobank.ac.uk/register-apply/. This research has been conducted using the UK Biobank Resource under Application Number 98358.

## 6. Acknowledgements

The authors express sincere gratitude to all individuals who generously participated in each one of the GWAS we have included in this study, as well as UK Biobank participants. This work uses data provided by patients and collected by the NHS as part of their care and support.

