## Supplementary Material for "PDE5 inhibition and Alzheimer’s disease risk: a mendelian randomisation study"

**Supplementary Note 1. Study details of the Alzheimer’s disease GWAS.**

**Meta-analysis of 74,046 individuals identifies 11 new susceptibility loci for Alzheimer’s disease, Lambert *et al*., 2013**

Study design: In stage 1, they used genotyped and imputed data (7,055,881 SNPs) to perform meta-analysis on 4 previously published GWAS data sets consisting of 17,008 Alzheimer's disease cases and 37,154 controls. In stage 2, 11,632 SNPs were genotyped and tested for association in an independent set of ~20,000 participants with ~40% of Alzheimer cases. All participants included in all the analysis were European.

Full description of the data sets used in stage 1 (I-GAP datasets):

1. **Alzheimer’s Disease Genetic Consortium (ADGC):** The ADGC dataset comprises subjects from:
   a. **The NIA ADC Samples (ADC):** The cohort included 2,288 autopsy-confirmed AD cases, 913 clinically confirmed AD cases, and 519 cognitively normal elders (CNEs) who where older than 60 years at age of death and 744 living CNEs. AD cases were demented according to DSM-IV criteria or Clinical Dementia Rating (CDR) ≥ 1. Persons with Down’s syndrome, non-AD tauopathies and synucleinopathies were excluded. Controls did not meet DSM-IV criteria for dementia, did not have a diagnosis of mild cognitive impairment (MCI), and had a CDR of 0, if performed. Controls that did not meet or were low-likelihood AD by NIA/Reagan criteria, had sparse or no amyloid plaques, and a Braak NFT stage of 0 – II.
   b. **Oregon Health and Science University (OHSU):** The dataset includes 132 autopsy-confirmed AD cases and 152 deceased controls that were evaluated for dementia within 12 months prior to death (age at death >65 years).
   c. **The ADNI Study (ADNI):** Longitudinal, multi-site observational study with 268 AD cases with MRI confirmation of AD diagnoses and 173 healthy controls with AD-free status confirmed as of most recent follow-up. AD subjects were between the ages of 55–90, had an MMSE score of 20–26 inclusive, met NINCDS/ADRDA criteria for probable AD, and had an MRI consistent with the diagnosis of AD. Control subjects had MMSE scores between 28 and 30 and a Clinical Dementia Rating of 0 without symptoms of depression, MCI or other dementia and no current use of psychoactive medications.
   d. **The MIRAGE Study (MIRAGE):** Family-based genetic epidemiological study of AD at 17 clinical centres in the United States, Canada, Germany and Greece. A total of 1,262 participants contributed, with 509 AD cases and 753 CNEs.
   e. **The NCRAD/NIA-LOAD Family Study (NCRAD/NIA-LOAD):** The study recruited families with two or more affected siblings with LOAD and unrelated. A total of 1,819 cases and 1,969 CNEs were recruited.
   f. **University of Miami/Vanderbilt University/Mt. Sinai School of Medicine (UM/VU/MSSM):** This dataset contains 1,186 cases and 1,135 CNEs ascertained at the University of Miami, Vanderbilt University and Mt. Sinai School of Medicine.
   g. **The ACT/eMERGE Studies (ACT):** The ACT cohort is an urban and suburban elderly population from a stable HMO. Initially, it enrolled 2,581 cognitively intact subjects ≥ 65, who were enrolled between 1994 and 1998, but additional subjects have enrolled until May 2009. In total, ACT/eMERGE contributed data on 566 individuals with probable or possible AD (70 with autopsy-confirmation) and on 1,696 CNEs (155 with autopsy-confirmation).
   h. **The GenADA Study:** GenADA cohort includes 669 AD cases and 713 CNEs ascertained from nine memory referral clinics in Canada between 2002 and 2005. All patients with AD satisfied NINCDS-ADRDA and DSM-IV criteria for probable AD with Global Deterioration Scale scores of 3-7. CNEs had MMSE test scores higher than 25 (mean 29.2 ± 1.1), a Mattis Dementia Rating Scale score of ≥ 136, a Clock Test without error, and no impairments on seven instrumental activities of daily living questions from the Duke Older American Resources and Services Procedures test.
   i. **The TGEN2 Study:** Among the TGEN2 data analysed were 864 clinically- and neuropathologically characterized brain donors, and 493 CNEs without dementia or significant AD pathology. Of these cases and CNEs, 667 were genotyped as a part of the TGEN1 series. Samples were obtained from twenty-one different National Institute on Aging-supported AD Centre brain banks. The criteria for inclusion were as follows: self-defined ethnicity of European descent, neuropathologically confirmed AD or neuropathology present at levels consistent with status as a control, and age of death greater than 65.
   j. **Mayo clinic:** All 728 cases and 1,173 controls consisted of Caucasian subjects from the United States ascertained at the Mayo Clinic. Subjects were diagnosed by a neurologist at the Mayo Clinic in Jacksonville, Florida or Rochester, Minnesota. The neurologist confirmed a Clinical Dementia Rating score of 0 for all controls; cases had diagnoses of possible or probable AD made according to NINCDS-ADRDA criteria. There were 221 autopsy-confirmed cases and 216 CNEs autopsy-confirmed controls. All AD brains analyzed in the study had a Braak score of 4.0 or greater. Brains employed as controls had a Braak score of 2.5 or lower but often had brain pathology unrelated to AD and pathological diagnoses that included vascular dementia, frontotemporal dementia, dementia with Lewy bodies, multi-system atrophy, amyotrophic lateral sclerosis, and progressive supranuclear palsy.
   k. **The ROS/MAP Studies:** Community-based cohort studies. The ROS has been ongoing since 1993, whereas the MAP since 1997. A total of 1,072 persons passed genotyping QC. Of these, 296 met clinical criteria for AD at the time of their last clinical evaluation or time of death and met neuropathologic criteria for AD for those on whom neuropathologic data were available, and 776 were without dementia or MCI at the time of their last clinical evaluation or time of death and did not meet neuropathologic criteria for AD for those on whom neuropathologic data were available.
   l. **University of Pittsburgh (UP):** The University of Pittsburgh dataset contains 1,271 European AD cases (of which 277 were autopsy-confirmed) recruited by the University of Pittsburgh Alzheimer’s Disease Research Centre, and 841 European, CNEs ages 60 and older (2 were autopsy-confirmed). All AD cases met NINCDS/ADRDA criteria for probable or definite AD.
   m. **Washington University (WU):** A European American LOAD case-control data-set consisting of 339 cases and 187 healthy elderly controls was used in analyses for this study.
2. **The Cohort for Heart and Ageing Research in Genomic Epidemiology (CHARGE) consortium**: The CHARGE Consortium encompasses six large, prospective, community-based cohort studies. However, two out of the six studies were excluded from the analysis as they had not systematically ascertained all their dementia cases at the time of the GWAS analysis. The remaining studies include:
   a. **The Cardiovascular Health Study CHS:** Prospective population-based cohort study of risk factors for vascular and metabolic disease that in 1989-90 enrolled adults aged ≥65 years, at the United States. Exclusion criteria included prevalent coronary artery disease, congestive heart failure, peripheral vascular disease, valvular heart disease, stroke, transient ischemic attack, dementia other than AD. The study sample was of 2,522. Cases were classified as: (1) Possible/probable AD without VaD (categorized as pure AD, included in all AD), (2) mixed AD (for cases that met criteria for both AD and VaD, included in all-AD), and (3) possible/probable VaD without AD (excluded from the present study).
   b. **The Framingham Heart Study (FHS):** Cohort study initiated in 1948 by recruiting adult population residing in Framingham, Massachusetts. Participants where followed-up Participants with vascular dementia were not disqualified from obtaining a concomitant diagnosis of AD.
   c. **The Rotterdam Study:** Participants from a district of Rotterdam (Ommoord) aged ≥55 years at the baseline examination in 1990-93. Participants were followed-up until 2004. Participants with dementia other than AD at baseline were excluded. Diagnoses of AD and VaD were not mutually exclusive.
   d. **Age, Gene/Environment Susceptibility-Reykjavik Study (AGES-RS):** Between 2002 and 2006, the study enrolled 5,764 participants. After randomised selection and genotyping quality control criteria, 2,807 were included in the GWAS and classified as either non-demented or having Alzheimer’s Disease (AD). 123 were diagnosed as dementia cases following rigorous criteria explained in the study. The 123 cases were further classified between: (1) AD without vascular dementia (VaD) (n = 55, included in the analysis), (2) mixed AD (n = 23, cases that met criteria for both AD and VaD, included in the analysis), (3) possible/probable VaD or other dementia without AD (n = 45, excluded for this study).
3. **European Alzheimer’s Disease Initiative (EADI) consortium:** 2,243 AD cases were ascertained by neurologists from Bordeaux, Dijon, Lille, Montpellier, Paris, and Rouen. Clinical diagnosis of probable AD was established according to the DSM-III-R and NINCDS-ADRDA criteria. Controls were selected from the 3C study, which is a population-based, prospective (7-years follow-up) study of the relationship between vascular factors and dementia. There were 9,294 subjects that agreed to participate and meet the inclusion criteria.
4. **Genetic and Environmental Risk in Alzheimer’s Disease (GERAD) consortium:** This consortium includes 3,177 AD cases and 7,277 controls from different studies. All AD cases met criteria for either probable (NINCDS-ADRDA, DSM-IV) or definite (CERAD) AD. All elderly controls were screened for dementia using the MMSE or ADAS-cog, were determined to be free from dementia at neuropathological examination or had a Braak score of 2.5 or lower

In stage 2, 22,618 case-control samples were obtained for replication from centres in Austria, Belgium, Finland, Germany, Greece, Hungary, Italy, Spain, Sweden, the UK and the USA. Clinical diagnoses of probable AD were all established according to the DSM-III-R and NINCDS-ADRDA criteria. Controls were defined as subjects without DMS-III-R dementia criteria and with integrity of their cognitive functions (MMS > 25). However, in this GWAS, stage 2 dataset is only used for validation and is not included in the main analysis.

### Common variants in Alzheimer’s disease and risk stratification by polygenic risk scores, de Rojas *et al*., 2021

Study design: Large genetic association study by merging all available case-control datasets and by-proxy study results (n = 409,435).

Full description of the data sets used: There were three AD GWASs involved: IGAP (30,344 AD cases and 52,427 controls, including stage 1 and 2), GR@ACE case-control study (6,331 AD cases and 6,055 controls) and UKB AD-by-proxy case-control study (27,696 cases of maternal AD with 250,980 controls, and 14,338 cases of paternal AD with 245,941 controls).

1. **I-GAP datasets**: Described carefully in Lambert *et al*. datasets description.
2. **GR@ACE study:** This study recruited AD patients from Fundació ACE, Institut Català de Neurociències Aplicades (Catalonia, Spain) and control individuals from across Spain^1^. Additional cases and controls were obtained from dementia cohorts included in the Dementia Genetics Spanish Consortium (DEGESCO)^2^. AD diagnosis was established by neurologists, neuropsychologists, and social workers according the DSM-IV criteria for dementia and National Institute on Aging and Alzheimer’s Association’s (NIA-AA) 2011 guidelines. AD cases were individuals with dementia diagnosed with probable or possible AD at any point in their clinical course.
3. **UK Biobank:** The UK Biobank is a prospective-cohort study that recruited over 500,000 participants aged 40-69 years when recruited in 2006-2010^3^. In this study, they used the published GWAS summary statistics of Marioni *et al.^4^*, where they use a proxy-AD approach. 27,696 participants whose mothers had dementia (maternal cases) were compared with the 260,980 participants whose mothers did not have dementia. Likewise, the 14,338 participants whose fathers had dementia (paternal cases) were compared with the 245,941 participants whose fathers did not have dementia.

### A genome-wide association study with 1,126,563 individuals identifies new risk loci for Alzheimer’s disease, Wightman *et al*., 2021

Study design: Genome-wide association study of LOAD with 90,338 (46,613 proxy) cases and 1,036,225 (318,246 proxy) controls.

### Full description of the data sets used: Description of the datasets meta-analysed:

### 1. DeCODE: This study included 7,002 Alzheimer’s patients and 181,572 controls. AD diagnoses was established according to NINCDS-ADRDA criteria. Controls were drawn for various research projects at deCODE Genetics.

### 2. UK Biobank: Previously described in Rojas *et al*., datasets description.

### 3. The Trøndelag Health Study (HUNT): It includes 1156 cases and 7,157 controls. Cases were defined as individuals diagnosed with ICD-10 G30.0 or F00*, or ICD-9 331.0 and controls were individuals last seen as healthy with no previous diagnosis of Alzheimer’s disease. All controls were more than 80 years old.

### 4. 23andMe: It consists of 3,807 cases and 359,839 controls. Among the controls, there were 19,638 individuals between the age of 45-60 and 340,201 individuals over 60. There were 130 cases between 45-60 and 3677 cases over the age of 60.

### 5. BioVU: It includes 600 cases and 36,059 controls. Cases were defined as individuals diagnosed with ICD-10 G30 and ICD-9 331.0. Controls were individuals without any of the following ICD-10 diagnoses; G30, F01, F02, F03, F10.27, F10.97, F13.27, F13.97, F18.17, F18.27, F18.97, F19.17, F19.27, F19.97, G31.0, G31.83 and the following ICD-9 diagnoses; 331.0 ,290, 291.2, 292.82, 294.1, 294.10, 294.11, 294.2, 294.20, 294.21, 331.19, 331.82. Individuals with a family history of dementia in their electronic health records were also excluded from the control sample.

### 6. DemGene, TwinGene, STSA, Gothenburg, and ANMerge: DemGene includes 1638 cases and 6059 controls, STSA 320 cases and 750 controls, and TwinGene 224 cases and 6321 controls. ANMerge data includes 366 cases and 259 controls. The Gothenburg H70 Birth Cohort Studies and Clinical AD from Sweden (Gothenburg) AD cases.

### 7. I-GAP: Described previously in Lambert *et al*., datasets description.

### 8. Finngen: Summary statistics for 1798 cases and 72,206 controls from Finngen. Cases were diagnosed with ICD-19 G301.

### 9. GR@CE: Previously described in Rojas *et al*., datasets description.

### New insights into the genetic etiology of Alzheimer’s disease and related dementias, Bellenguez *et al*., 2022

Study design: Two-stage genome-wide association study totalling 111,326 clinically diagnosed/”proxy” AD cases and 677,663 controls.

### Full description of the data sets used: Stage 1 datasets are described below.

### 1. The European Alzheimer & Dementia Biobank dataset (EADB): The consortium groups together 20,464 Alzheimer’s disease (AD) cases and 22,244 controls after quality controls from 15 European countries.

### a. EADB-France: In the France node, samples were collected from nine countries and included 13,867 AD cases and 15,310 controls.

### b. EADB-Germany: In the German node, samples were collected from seven countries and obtained 4,159 AD cases and 4,545 controls.

### c. EADB-Netherlands: In the Dutch node, samples were collected from six organizations in the Netherlands and included 2,438 AD cases and 2,389 controls.

### d. EADB-Australia: It only included the Sydney MAS study, which accounts of 43 AD cases and 215 controls. Due to its sample size, it was not included in the meta-analysis.

### 2. GR@ACE study: Previously described in Rojas *et al*., datasets description.

### 3. The Rotterdam study: Previously described in Lambert *et al*., datasets description.

### 4. The European Alzheimer’s Disease Initiative (EADI) Consortium: Previously described in Lambert *et al*., datasets description.

### 5. Genetic and Environmental Risk in AD (GERAD) Consortium/Defining Genetic, Polygenic, and Environmental Risk for Alzheimer’s Disease (PERADES) Consortium: Previously described in Lambert *et al*., datasets description.

### 6. The Norwegian DemGene Network: It includes clinical sites with 2,225 cases and 3,089 healthy controls from different studies. Cases were diagnosed according the NINCDS-ADRDA criteria or the ICD-10 research criteria.

### 7. The Neocodex-Murcia study (NxC): It includes 324 sporadic AD patients and 754 controls of unknown cognitive status from the Spanish general population. A D patients were diagnosed as having possible or probable AD in accordance with the NINCDS–ADRDA criteria.

### 8. The Copenhagen City Heart Study (CCHS): CCHS is a prospective study of the Danish general population. Individuals were selected randomly based on the national Danish Civil Registration System.

### 9. Bonn studies: It includes DietBB sample and the Bonn OMNI cohort. In the first one, participants were recruited in six German cities. AD cases were diagnosed according to the criteria set of DSM-IV and NINCDS-ADRDA criteria. Mixed dementia was diagnosed in cases of cerebrovascular events without temporal relationship to cognitive decline. Mixed dementia and dementia in AD were combined. The Bonn OMNI cohort consists of 1,095 patients with mild cognitive impairment (MCI) and 648 cases with mild Alzheimer’s disease (AD) clinical dementia syndrome

### 10. UK Biobank: Previously described in Rojas *et al*., datasets description.

**Supplementary Table 1.** Summary of the Alzheimer’s disease GWAS used in the main analysis (Lambert *et al.*) and in the secondary analysis (De Rojas *et al.,* Wightman *et al.,* and Bellenguez *et al.*).

| **Year** | **Author** | **Consortium** | **Total N** | **Cases (%)** | **Mean age at assessment^a^** | | **SNP-based heritability on liability scale (5% prevalence)** | **GWAS assembly** | **Ancestry** | **Proxy cases** | **Link** |
| --- | --- | --- | --- | --- | --- | --- | --- | --- | --- | --- | --- |
|  |  |  |  |  | **Cases** | **Controls** |  |  |  |  |  |
| 2013 | Lambert *et al.* | IGAP | 54,162 | 17,008 (31) | 76.6 | 70.5 | 0.09 (0.02) | hg19/  GRCh37 | European | No | https://ftp.ebi.ac.uk/pub/databases/gwas/summary_statistics/GCST002001-GCST003000/GCST002245/ |
| 2021 | De Rojas *et al.* | IGAP,  Gr@ce,  UK Biobank | 409,435 | 81,611  (20) | Not known | 67.3 | 0.03 (0.004) | hg38/  GRCh38 | European | Yes | <https://fundacioace-my.sharepoint.com/personal/iderojas_fundacioace_org/_layouts/15/onedrive.aspx?id=%2Fpersonal%2Fiderojas%5Ffundacioace%5Forg%2FDocuments%2FPRS%2Fgrace%5Fstg1%262%2FPAPER%20DRAFT%2F04%5FNatCom%2FmetaGWAS%5FDiscovery%5Fdata%5Fshared%2FSumstats%5FSPIGAPUK2%5F20190625%2Ezip&parent=%2Fpersonal%2Fiderojas%5Ffundacioace%5Forg%2FDocuments%2FPRS%2Fgrace%5Fstg1%262%2FPAPER%20DRAFT%2F04%5FNatCom%2FmetaGWAS%5FDiscovery%5Fdata%5Fshared&ga=1> |
| 2021 | Wightman *et al.* | IGAP  GR@ACE  …  (It excludes UK Biobank and 23 and ME) | 398,108 | 39,968  (10) | NA | NA | 0.03  (0.006)^b^ | hg19/  GRCh37 | European | No | <https://ftp.ebi.ac.uk/pub/databases/gwas/summary_statistics/GCST013001-GCST014000/GCST013196/> |
| 2022 | Bellenguez *et al*. | IGAP,  GR@ACE,  UK Biobank,  … | 487,511 | 85,934 (18) | 67.2 | 57.9 | 0.03 (0.003) | hg38/  GRCh38 | European | Yes | <https://ftp.ebi.ac.uk/pub/databases/gwas/summary_statistics/GCST90027001-GCST90028000/GCST90027158/> |

^a^Mean age at assessment. If not reported, was estimated as weighted (by sample sizes) average of the ages at assessments reported in the contributed studies.

^b^Calculated without UK Biobank

**Supplementary Table 2.** UK Biobank codes.

| **Description** | **Field** |
| --- | --- |
| ***Baseline characteristics*** |  |
| Sex | 31 |
| Age at first assessment | 21003 |
| Ethnic background | 21000 |
| Body mass index | 21001 |
| ***Alzheimer’s disease cohort*** | |
| Date of dementia report | 42018 |
| Date of vascular dementia report | 42022 |
| Date of frontotemporal dementia report | 42024 |
| Date of Alzheimer’s disease report | 42020 |
| ***Regression*** |  |
| Principal components | 22009.0.1-22009.0.10 |
| Genetic batch | 22000 |

**Supplementary Table 3. Linkage disequilibrium matrix of the instruments.** Notice that values correspond to r, not r^2^.

| **SNP** | **rs10050092 (C/T)** | **rs12646525 (T/C)** | **rs17355550 (C/T)** | **rs66887589 (C/T)** | **rs80223330 (A/G)** |
| --- | --- | --- | --- | --- | --- |
| **rs10050092 (C/T)** | 1.00 | -0.24 | -0.17 | -0.64 | -0.32 |
| **rs12646525 (T/C)** | -0.24 | 1.00 | 0.36 | -0.47 | -0.18 |
| **rs17355550 (C/T)** | -0.17 | 0.36 | 1.00 | -0.13 | -0.07 |
| **rs66887589 (C/T)** | -0.64 | -0.47 | -0.13 | 1.00 | 0.38 |
| **rs80223330 (A/G)** | -0.32 | -0.18 | -0.07 | 0.38 | 1.00 |

**Supplementary Table 4. Harmonised variants respect the alleles of the linkage disequilibrium matrix.** Note: EA = Effect allele, OA = Other allele, SE = Standard error, EAF = Effect allele frequency.

| **Study outcome** | **SNP** | **EA** | **OA** | **EAF** | | **Beta** | | **SE** | | | **P-Value** | | |
| --- | --- | --- | --- | --- | --- | --- | --- | --- | --- | --- | --- | --- | --- |
|  |  |  |  | **Exposure** | **Outcome** | **Exposure** | **Outcome** | | **Exposure** | **Outcome** | | **Exposure** | **Outcome** |
| **Lambert *et al*. (2013)** | rs17355550 | C | T | 0.03 |  | -0.14 | 0.10 | | 0.05 | 0.05 | | 4.1e-03 | 5.3e-02 |
|  | rs80223330 | A | G | 0.14 |  | 0.10 | 0.04 | | 0.03 | 0.03 | | 1.2e-04 | 2.0e-01 |
|  | rs12646525 | T | C | 0.22 |  | -0.10 | 0.00 | | 0.02 | 0.02 | | 6.7e-06 | 9.9e-01 |
|  | rs66887589 | C | T | 0.48 |  | 0.16 | 0.00 | | 0.02 | 0.02 | | 1.8e-20 | 8.8e-01 |
|  | rs10050092 | C | T | 0.34 |  | -0.13 | -0.01 | | 0.02 | 0.02 | | 8.6e-13 | 6.1e-01 |
| **Wightman *et al*. (2021)** | rs17355550 | C | T | 0.03 | 0.04 | -0.14 | 0.04 | | 0.05 | 0.03 | | 4.1e-03 | 1.1e-01 |
|  | rs80223330 | A | G | 0.14 | 0.16 | 0.10 | 0.00 | | 0.03 | 0.01 | | 1.2e-04 | 9.9e-01 |
|  | rs12646525 | T | C | 0.22 | 0.25 | -0.10 | 0.02 | | 0.02 | 0.01 | | 6.7e-06 | 1.2e-01 |
|  | rs66887589 | C | T | 0.48 | 0.46 | 0.16 | -0.01 | | 0.02 | 0.01 | | 1.8e-20 | 2.0e-01 |
|  | rs10050092 | C | T | 0.34 | 0.32 | -0.13 | 0.00 | | 0.02 | 0.01 | | 8.6e-13 | 7.7e-01 |
| **De Rojas *et al*. (2021)** | rs17355550 | C | T | 0.03 |  | -0.14 | 0.03 | | 0.05 | 0.03 | | 4.1e-03 | 3.2e-01 |
|  | rs80223330 | A | G | 0.14 |  | 0.10 | 0.01 | | 0.03 | 0.01 | | 1.2e-04 | 4.8e-01 |
|  | rs12646525 | T | C | 0.22 |  | -0.10 | 0.00 | | 0.02 | 0.01 | | 6.7e-06 | 9.0e-01 |
|  | rs66887589 | C | T | 0.48 |  | 0.16 | 0.00 | | 0.02 | 0.01 | | 1.8e-20 | 9.7e-01 |
|  | rs10050092 | C | T | 0.34 |  | -0.13 | 0.00 | | 0.02 | 0.01 | | 8.6e-13 | 9.9e-01 |
| **Bellenguez *et al*. (2022)** | rs17355550 | C | T | 0.03 | 0.03 | -0.14 | 0.00 | | 0.05 | 0.02 | | 4.1e-03 | 8.6e-01 |
|  | rs80223330 | A | G | 0.14 | 0.14 | 0.10 | 0.01 | | 0.03 | 0.01 | | 1.2e-04 | 5.5e-01 |
|  | rs12646525 | T | C | 0.22 | 0.22 | -0.10 | 0.00 | | 0.02 | 0.01 | | 6.7e-06 | 9.8e-01 |
|  | rs66887589 | C | T | 0.48 | 0.48 | 0.16 | 0.00 | | 0.02 | 0.01 | | 1.8e-20 | 6.1e-01 |
|  | rs10050092 | C | T | 0.34 | 0.33 | -0.13 | 0.00 | | 0.02 | 0.01 | | 8.6e-13 | 7.8e-01 |

**Supplementary Table 5. Baseline characteristics of the sex-specific cohorts.** Note: IMD = Index of Multiple Deprivation, SD = Standard deviance, AD = Alzheimer’s disease.

| **Variable** | **Males** | | **Females** | |
| --- | --- | --- | --- | --- |
|  | **AD Controls** | **AD Cases** | **AD Controls** | **AD Cases** |
| N (%) | 219,063 (99.4) | 1,289 (0.6) | 260,575 (99.4) | 1,462 (0.6) |
| Age at first assessment (Mean (SD)) | 56.61 (8.18) | 64.75 (4.35) | 56.26 (7.98) | 64.57 (4.21) |
| IMD (Mean (SD)) | 17.57 (14.33) | 18.67 (14.81) | 16.97 (13.73) | 18.30 (14.53) |
| Ethnic background (%) |  |  |  |  |
| White | 198,743 (99.4) | 1,199 (0.6) | 235,551 (99.4) | 1,355 (0.6) |
| Non-white | 19,331 (99.6) | 86 (0.4) | 24,262 (99.6) | 100 (0.4) |
| Unknown | 989 (99.6) | 4 (0.4) | 762 (99.1) | 7 (0.9) |
| Body mass index (Mean (SD)) | 27.83 (4.23) | 27.65 (4.24) | 27.06 (5.18) | 27.13 (5.13) |

**Supplementary Table 6. Two-Step Mendelian randomisation results.** Note: OR = Odds ratio, SE = Standard Error, SNPs = Single Nucleotide Polymorphism, GWAS = Genome wide association study, BMI = Body mass index,

| **Study outcome** | **Confounder** | **GWAS ID** | **OR** | **SE** | **P-Value** |
| --- | --- | --- | --- | --- | --- |
| Lambert *et al*. (2013) | BMI | ukb-b-19953 | 0.99 | 0.11 | 0.96 |
| Wightman *et al*. (2021) | BMI | ukb-b-19953 | 0.91 | 0.06 | 0.10 |
| De Rojas *et al*. (2021) | BMI | ukb-b-19953 | 0.98 | 0.05 | 0.74 |
| Bellenguez *et al*. (2022) | BMI | ukb-b-19953 | 1.01 | 0.05 | 0.89 |

**Supplementary Table 7. Leave-one-out analysis results.** Note: SNP = Single Nucleotide Polymorphism, OR = Odds ratio, SE = Standard Error.

| **Study outcome** | **SNP** | **N Instruments** | **OR** | **SE** | **P-Value** |
| --- | --- | --- | --- | --- | --- |
| Lambert *et al*. (2013) | All | 5 | 1.001 | 0.02 | 0.96 |
| Lambert *et al*. (2013) | rs10050092 | 4 | 1.003 | 0.03 | 0.90 |
| Lambert *et al*. (2013) | rs66887589 | 4 | 1.000 | 0.02 | 1.00 |
| Lambert *et al*. (2013) | rs12646525 | 4 | 1.004 | 0.02 | 0.88 |
| Lambert *et al*. (2013) | rs80223330 | 4 | 0.999 | 0.02 | 0.98 |
| Lambert *et al*. (2013) | rs17355550 | 4 | 0.995 | 0.02 | 0.77 |
| Wightman *et al*. (2021) | All | 5 | 1.016 | 0.01 | 0.11 |
| Wightman *et al*. (2021) | rs10050092 | 4 | 1.016 | 0.01 | 0.13 |
| Wightman *et al*. (2021) | rs66887589 | 4 | 1.017 | 0.01 | 0.11 |
| Wightman *et al*. (2021) | rs12646525 | 4 | 1.015 | 0.01 | 0.14 |
| Wightman *et al*. (2021) | rs80223330 | 4 | 1.016 | 0.01 | 0.11 |
| Wightman *et al*. (2021) | rs17355550 | 4 | 1.014 | 0.01 | 0.18 |
| De Rojas *et al*. (2021) | All | 5 | 1.003 | 0.01 | 0.76 |
| De Rojas *et al*. (2021) | rs10050092 | 4 | 1.002 | 0.01 | 0.84 |
| De Rojas *et al*. (2021) | rs66887589 | 4 | 1.003 | 0.01 | 0.75 |
| De Rojas *et al*. (2021) | rs12646525 | 4 | 1.003 | 0.01 | 0.78 |
| De Rojas *et al*. (2021) | rs80223330 | 4 | 1.003 | 0.01 | 0.77 |
| De Rojas *et al*. (2021) | rs17355550 | 4 | 1.001 | 0.01 | 0.94 |
| Bellenguez *et al*. (2022) | All | 5 | 0.997 | 0.01 | 0.77 |
| Bellenguez *et al*. (2022) | rs10050092 | 4 | 0.996 | 0.01 | 0.62 |
| Bellenguez *et al*. (2022) | rs66887589 | 4 | 0.998 | 0.01 | 0.82 |
| Bellenguez *et al*. (2022) | rs12646525 | 4 | 0.996 | 0.01 | 0.67 |
| Bellenguez *et al*. (2022) | rs80223330 | 4 | 0.998 | 0.01 | 0.77 |
| Bellenguez *et al*. (2022) | rs17355550 | 4 | 0.997 | 0.01 | 0.72 |

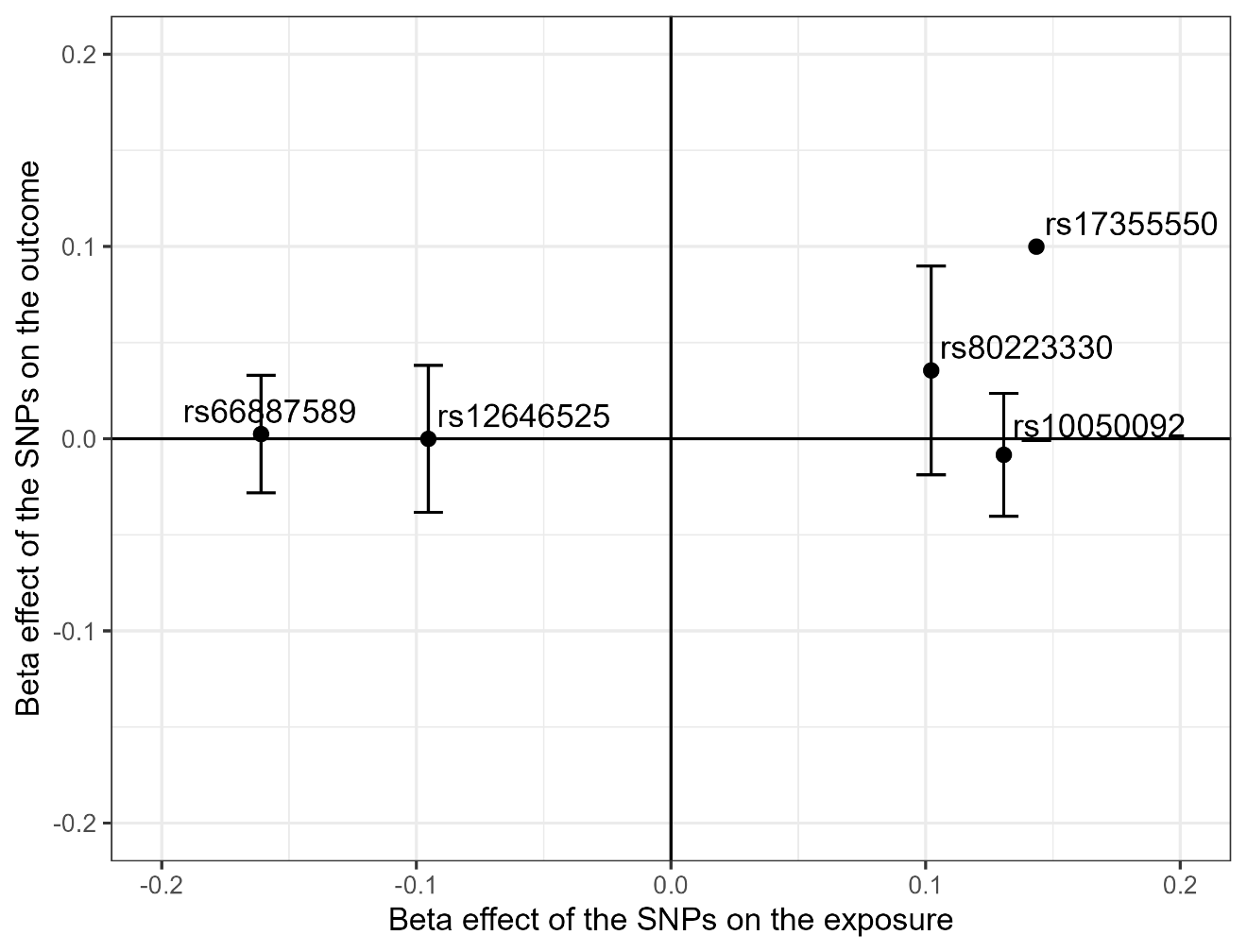

**Supplementary Fig. 1: SNPs effect on the exposure against SNPs effect on the outcome (using Lambert’s *et al.* GWAS for the outcome).**

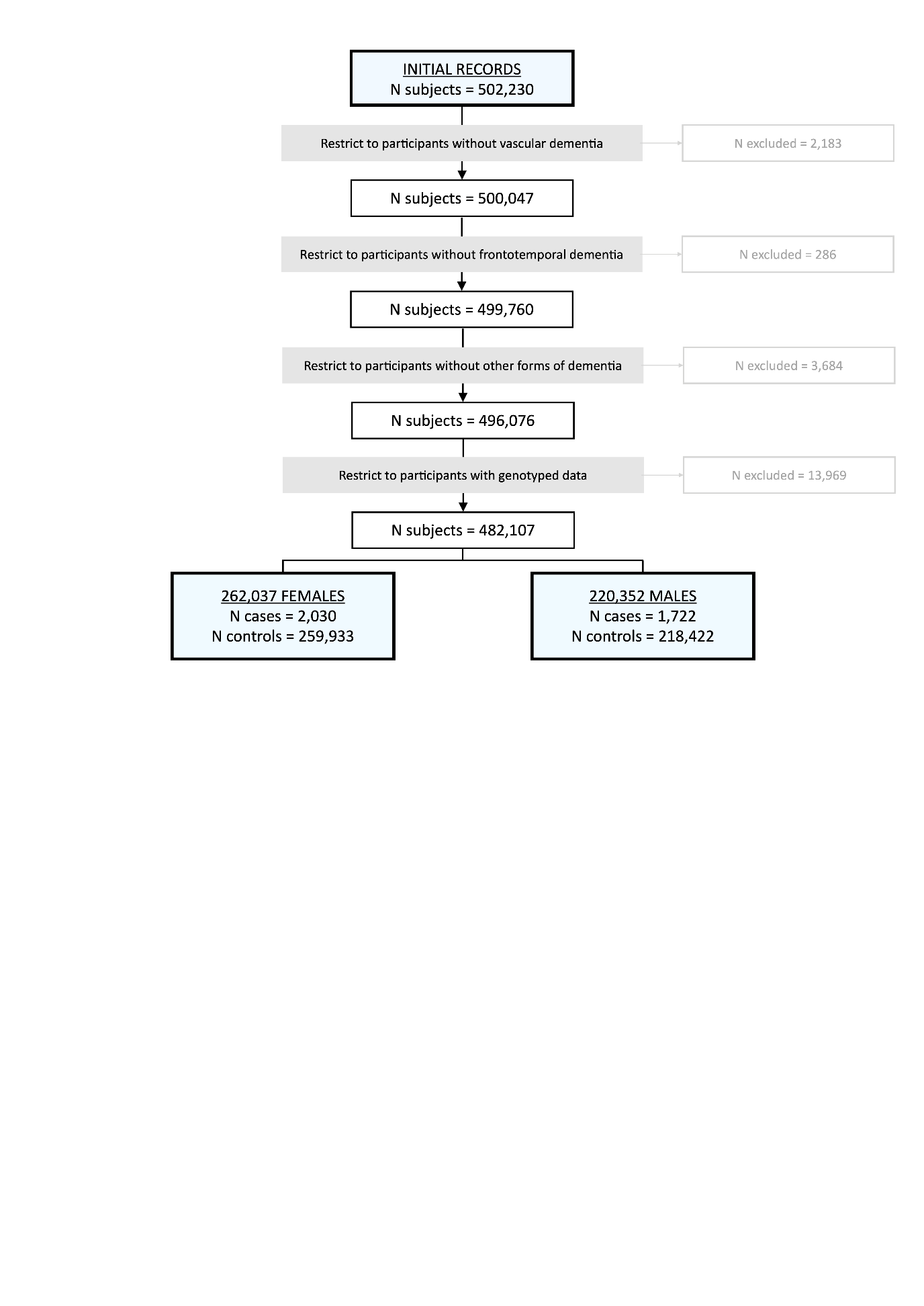

**Supplementary Fig. 2: UK Biobank flowchart.**

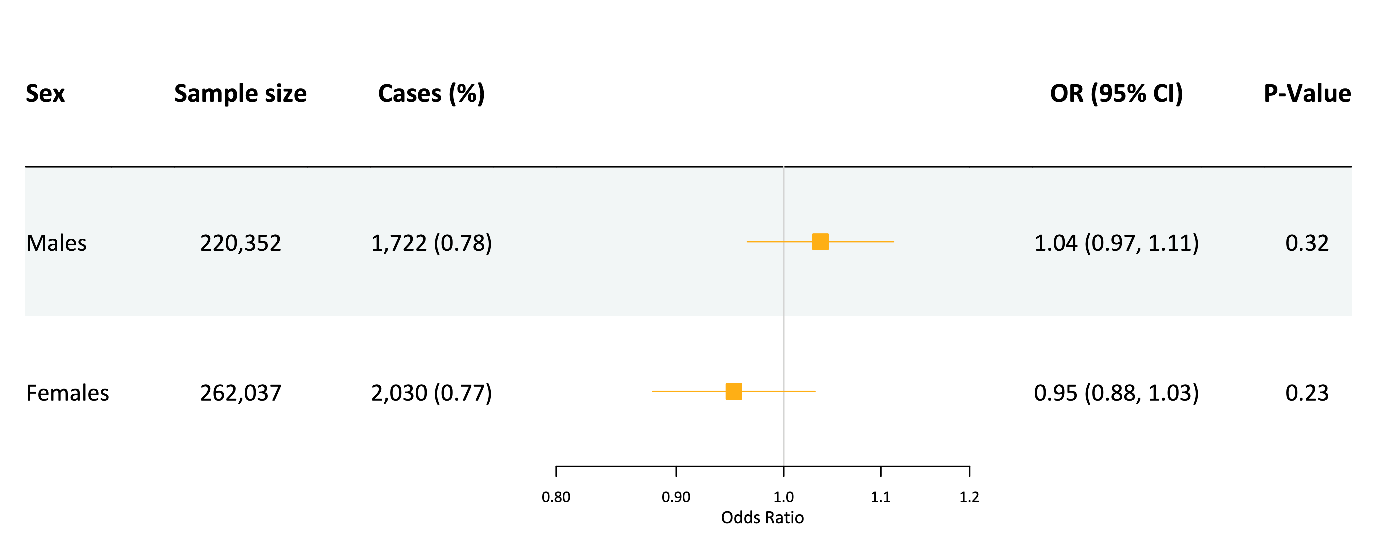

**Supplementary Fig. 3: Mendelian randomisation results of the sex-specific analysis.** Odds ratio is scaled to represent 100mg dose of sildenafil (which reduces 5mmHg diastolic blood pressure). Note: OR = Odds ratio.
